# *Laga* Ecosystem, Species Entanglements and the Risk of Zoonotic Disease Transmission: A Multi-Site, Multi-Method Ethnographic Study

**DOI:** 10.1101/2024.05.23.24307773

**Authors:** Dalmas Omia, Dismas Oketch, Ruth Njoroge, Isaac Ngere, John Gachohi, Samuel Waiguru, Abdulai G. Magarre, Samoel Ashimosi Khamadi, Scott L Nuismer, John Mwaniki Njeru, Boku Bodha, Nazaria Nyaga, Humphrey Njaanake, Walter Jaoko, Kariuki Njenga, Eric Osoro

## Abstract

Dry riverbeds, also called *Iaga*, are a complex ecosystem of multispecies interactions between livestock, humans, microorganisms, and their environment. Despite *laga’s* One Health entanglement of species and environment, few studies have explored the risks of transmission of diseases through direct herd-herd or herd-human contact or indirect contact with fomites surrounding the *laga*. This study focuses on ethnographic and epidemiological investigations on *lagas* within Kenya. The study deploys qualitative multimethod-walking interviews, in-depth interviews, key informant interviews, focus group discussions and observations to collect the data from Marsabit and Kajiado Counties in Kenya. Results point to the comingling of infected and healthy herds, cross-livestock species mixing, sharing of watering troughs, and feeding dogs placental and parturition materials at the herd level. The human transmission risks include non-protective parturition assistance, the use of camel urine as an antiseptic substance, humans sharing animal-watering troughs, and consuming non-processed milk. Further, the fomites comprise contaminated excreta, infected placental materials on *laga* stones, deposition of infected aborted fetuses on the *laga* body, and bacteria in the sand that end up ingested or inhaled as dust during dry seasons. The study concludes that intensified water insecurity due to climate variability will deepen multispecies interactions at the *laga* given that it holds a lifeline in drylands for pastoralists, hence, heightening brucellosis transmission risks. The study’s results recommend a reinvention of brucellosis preventive measures that consider the pathogen flux within *laga* systems and multispecies interactions. Such an approach should consider the multidimensional-clinical, environmental, and cultural co-production of solutions where preventive behaviors are prioritized.

## 1. Introduction

Brucellosis is an endemic zoonotic disease that remains neglected in most of the developing world where it causes devastating losses to the livestock industry and public health (Franc et al.,2018). The Bacteria that causes brucellosis belongs to the genus *Brucella* (Djangwani *et al*., 2021; Godfroid *et al*., 2014) of which the most relevant species to livestock health and public health are *Brucella abortus, Brucella melitensis, Brucella ovis* and *Brucella suis* known to infect cattle, small ruminants, and swine respectively (Djangwani *et al*., 2021; Osoro et al., 2015) Brucella ovis in sheep (Matle et al., 2021) and *Brucella canis* in dogs (Olsen & Palmer, 2014).

*Brucella* transmission occurs between animals and also between animals and humans (Lokamar *et al*., 2022; Djangwani *et al*., 2021; Njeru *et al*., 2016). Animals are thought to be infected primarily by consuming contaminated pasture or water, aborted foetuses, and foetal membranes, but may also be directly infected through contact with the genitalia of infected animals by licking (El-Sayed & Awad, 2018). Additionally, infected males can transmit the infection to females through natural mating and artificial insemination (Ragan 2021). Humans are generally infected through consumption of unpasteurized milk, or direct contact with the placenta, fetus, fetal fluids, and vaginal secretions of infected animals (Godfroid et al., 2011). Human-to-human transmission of brucellosis has been characterized in rare instances such as infected mothers breastfeeding their infants in blood transfusions and through sexual encounter (Tuon, Gondolfo & Cerchiari, 2017; CDC, 2019; Melzer et al., 2010).

In most cases, infection typically happens when the pathogen is ingested or enters the body through mucous membranes. However, *B. abortus* can also be transmitted through damaged or broken skin and through contaminated objects and surfaces (fomites). *Brucellae* can persist in the environment or soil, particularly under conditions of elevated humidity, cold temperatures, and minimal sun exposure. Moreover, it can maintain its viability for several months in water, aborted fetuses, and excreta, provided that the conditions are suitable (Abubakar, Mansoor & Arshed, 2012).

Brucellosis causes abortions and other reproductive disorders such as stillbirths, weak calves, retained placenta, and longer calving intervals in female animals (Djangwani et al., 2021; Njeru et al., 2016), while in humans, it results in a febrile illness characterized by intermittent fevers, sweats, chills, weakness, malaise, headache, anorexia, joint, and muscle pain (Djangwani et al., 2021; Food and Drug Administration, 2012; Pappal et al., 2006). Although Brucellosis has been largely controlled in high-income countries (Rubach et al., 2013; Osoro et al., 2015; Frank et al., 2018), it remains a persistent threat in many Low and Middle-Income countries (Frank et al., 2018; WHO, 2015; Rubach et al., 2013). For instance, El-Sayed and Awad (2018) note over 500,000 new cases of brucellosis annually, and Seleem et al. (2010) suggest the disease may be spreading into new areas and continually resurfacing within its historical range. Djangwani et al. (2021) report brucellosis prevalence in East African Community (EAC) countries, mostly in cattle (from 0.2% to 44%), with limited studies on small ruminants indicating prevalence in goats (between 0.0% and 20%) and sheep (up to 14%). In Kenya, brucellosis is prevalent among pastoralist communities ((Mwatondo et al., 2023; Lokamar et al., 2022; Djangwani et al., 2021;) and closely related to their mode of life (Osoro et al., 2015) and practices (Njenga et al., 2020).

Despite the overwhelming interest in brucellosis from biomedical and veterinary sciences, few studies have examined the transmission risks within the *laga* ecosystem which form the arteries of pastoralism in drylands with significant implications for One Health (Adisasmito et al., 2022). The *laga* plays a significant role in the daily lives of the community, serving as a natural and cultural point of interaction for all living organisms in arid regions. It is a source of water for drinking and bathing, a critical resource for watering livestock, and a meeting point for restocking food for families left behind and those leaving for grazing lands (*fora*). These close interactions typing species entanglements coupled with multiple practices within the *laga* are significant in OH, especially, in identifying possible risks of *Brucella* transmission by unpacking human-animal, animal-fomite, and animal-animal interactions.

We note that while the *laga* is a critical lifeline for the pastoral communities and their livestock, it has often been overlooked by epidemiological studies despite being a melting point for humans, animals and microorganisms. Consequently, taking the OH approach generates an understanding of how the interactions occur, enables contextualisation and reveals previously unknown opportunities in Brucellosis prevention and control strategies in drylands. Therefore, the study enriches the existing epidemiological evidence on the transmission pathways of *Brucella* by projecting the perspectives and experiences of local communities using ethnographic research method. This approach enhances the emergence of ideas from the community as opposed to the researchers imposing their ethnocentric meanings, hence, a very empowering method of inquiry. Additionally, the variations in these methods and their complementarity help to unpack the embedded reality of Brucella transmission risks. Furthermore, the use of multiple methods as part of the ethnographic approach expands the methodological toolbox of anthropology, demonstrating that diversity in approach can uncover greater depth of data. These approaches are significantly inductive and are important in informing ‘what’ and ‘where’ to sample for clinical work.

Our results are organised as follows: first, we begin with herd-to-herd transmission risks followed by herd-to-human and domestic-wild herbivores. We conclude by examining the environmental risk factors associated with fomites on the *laga*.

## 2 Materials and methods

### 2.1 Study areas

The study was conducted in two sites located in Kajiado and Marsabit

**Kajiado County** is located in the Rift Valley of Kenya between longitudes 36^0^ 5’ and 37^0^ 5’ east and between latitudes 10^0^’ and 30^0^’ south (NDMA, 2023). The county occupies 21,902 square kilometres with 1,268,261 people (KNBS, 2022) and the estimated distribution by livelihoods as; 42% pastoralists, 35% in formal employment or casual labour, 12% being agro-pastoralists and eight% deriving their livelihood from mixed farming (Figure 1). The study was conducted within Mailua-2° 19’ 22” S and longitude 36° 54’ 59” E. The maasai community in Mailua, among whom the current study was conducted, is largely involved in pastoralism as the main economic activity with a small percentage engaged in small-scale rainfed and irrigated crop farming.

**Figure 1.**
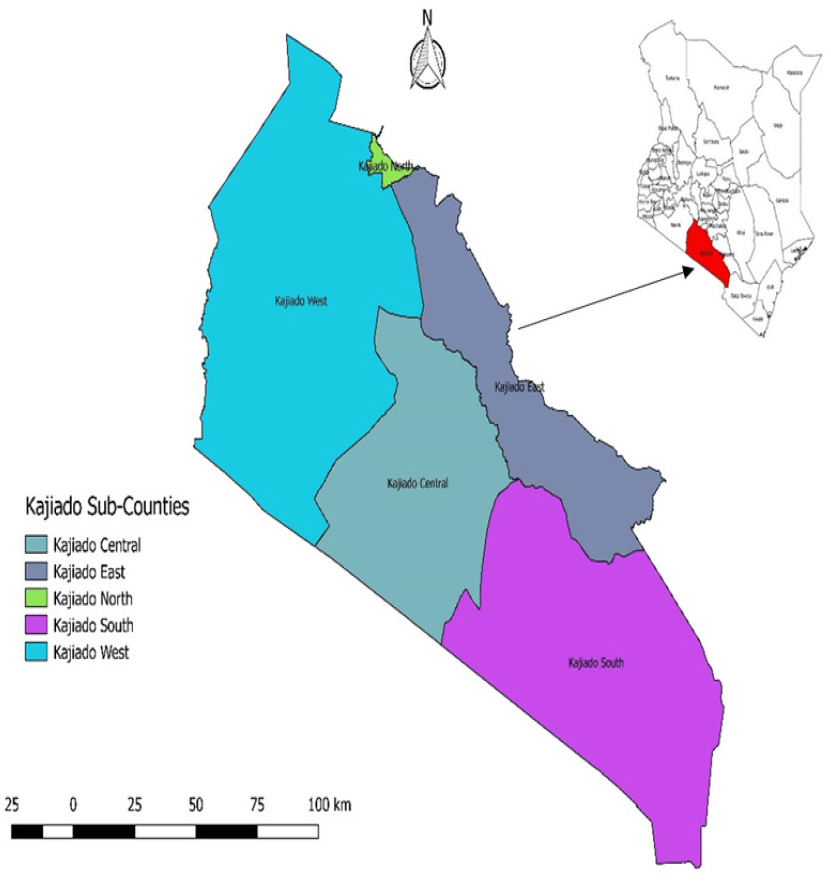
Kajiado County Source: NDMA (2023)

**Marsabit County** is located in the upper eastern region of Kenya and covers an area of 70,961.2km^2^ with a population of 515,000. It lies between latitude 10 58’N and 20 1’ S and longitude 38^0^ 34’E and 410 32’E (KNBS, 2022). The county has three main livelihood zones which include the pastoral livelihood zone constituting 81 per cent of the county population (Figure 2), the agro-pastoral livelihood zone comprising 16 % of the population and others having a combined population of 3% (IEBC, 2012).

**Figure 2.**
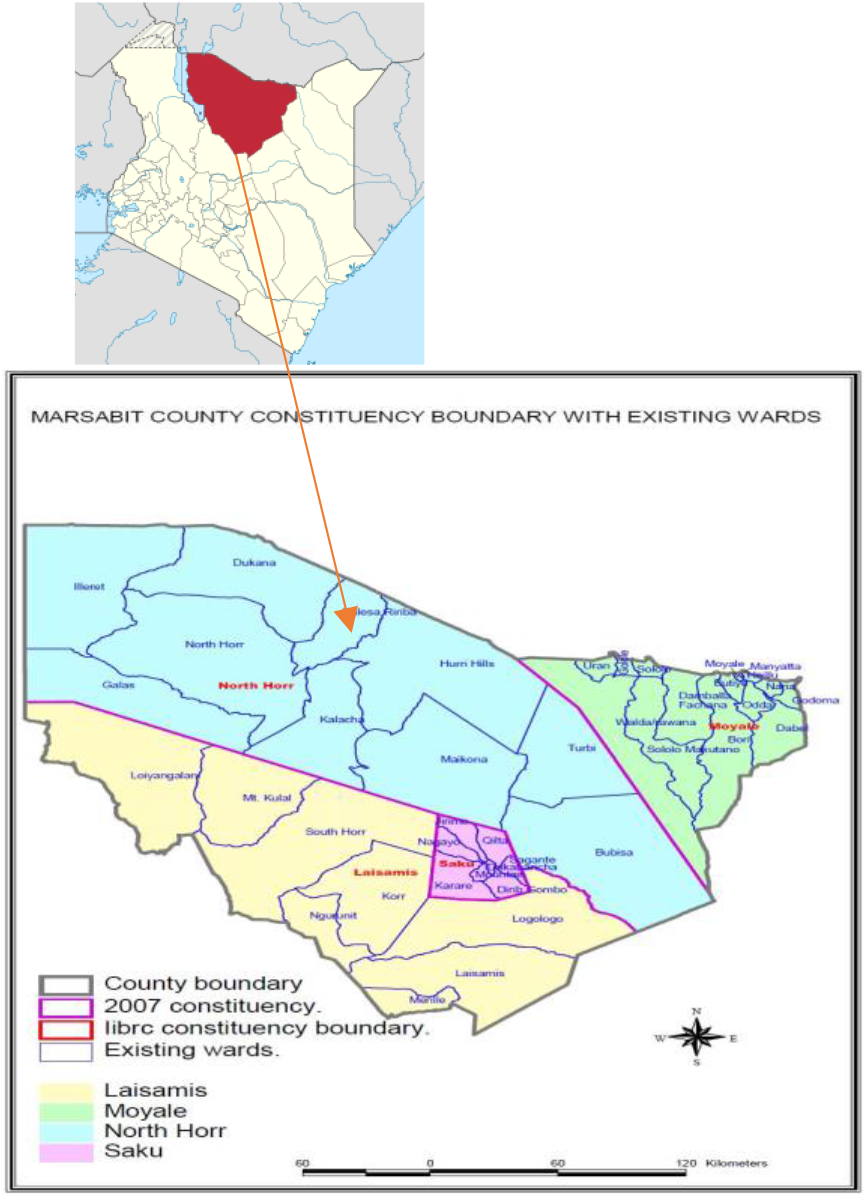
Marsabit County, Source: IEBC (2012)

The study was conducted within Laisamis Sub-County which lies between longitude 36 400 east and Latitude 00 150 south within the latitude 02° 450 north and 04° 27° north and longitude 37° 57° east and 39° 21° east. In particular, it was conducted among the Rendile community living within ecological zones V and VI of Laisamis including the lower slopes of volcanic and basement piles lying between 700 and 1,000m. Additionally, it covered extremely dry areas characterized as “bushed stone land,” comprising all hills and plains below 700m with typical dwarf-shrub grassland or a very dry form of bushed grassland. Rendile are camel herders due to the minimal rainfall received annually of about 200mm to 1000mm (Fratkin et al., 2004).

### 2.2 Ethical approval, consenting and participation

The Kenya Medical Research Institute Scientific and Ethics Review Unit reviewed and approved this study and assigned it approval number 4405. The study also obtained approval from Washington State University Institutional Review Board (IRB) and the National Commission for Science, Technology and Innovation (NACOSTI/P/22/17621). To enhance best practices in human-related research, the research team took a self-paced training on Good Clinical Practice research ethics and the protection of human subjects in research. Further, pre-field work ethical training and simulation exercises were carried out to build the capacities of the research team.

The research team obtained written, informed consent from study informants and participants before their engagement. A literate witness known to the potential informant was called upon to explain the nature, risks and benefits of the study to respondents who could not read and write. The research team gathered demographic information of the informants to understand their backgrounds as active pastoralists who are at risk of contracting brucellosis. The informants and participants received KES 1,000 (approx.US$10) as compensation for their time.

### 2.3 Study design, sampling and data collection

This was an exploratory, multi-method, multi-site ethnographic study in two pastoral communities examining Brucella transmission pathways within the *laga*. The research team limited the sampling to communities within a 20-30 km radius of the study reference health facility. This was the health facility that served majority of the inhabitants of the study catchment area. The study team was deliberate in picking *laga* sites where communities water their animals in large numbers and with diverse species of livestock.

The study employed interviews which took place in two phases. At first, study researchers held informal conversations with local herdsmen in the *laga* space during preliminary visits. At the same time, direct observations of the *laga* environment with its rich ecosystems were made and captured by way of photography. Walking interviews (Anderson, 2009; Bergeron et al., 2014), aimed at understanding how movement and space influence the perspectives of the informants, build a grounded understanding of the value of the *laga* to the livestock and community at large based on their everyday practices.

In the second phase, the qualitative research team, along with community health providers, conducted interviews with purposively drawn participants. These included in-depth interviews (n=60) and 20 Focus Group Discussions (n=198) with local users-pastoralists-aimed at building detailed narratives of *laga* ecosystems. Following this, key informant interviews (n=15) were conducted. Informal interviews at the *laga* were captured via descriptive notes, while the IDIs and FGDs were captured via audio recorders for verbatim transcription.

### 2.4 Data storage and protection

Data were stored in the Washington State University’s (WSU) One-Drive which includes encryption at rest and backed up in a hard drive accessible to the principal investigator. Only the research staff specifically involved in the qualitative data collection and management were granted access to allow for the anonymization and de-identification of the original audio files and transcripts. Even then, individual identifiers were not used as each participant was assigned a unique code number.

### 2.5 Data analysis

Debriefing sessions were held at the close of every fieldwork day to get a summary of key issues and emerging issues in each FGD, KII and IDI conversation. These initial reflections helped to contextualize some of the observed activities, and interactions between humans and livestock at the *laga*. The exercises helped to establish the emerging themes of the study following collective insights on the value and nature of observed behaviours including reflections on photographs taken. Each week of fieldwork was followed by a buffer day – a day when RAs were expected to transcribe the audio, translate them into English, type them out, and finally, check for clarity and completeness of the verbatim notes, before submitting them to the lead ethnographer. Subsequently, NVivo 14 (version 14.23.2) was used to organize, code, and disaggregate the textual material for qualitative analysis. Data was analyzed deductively based on themes that emerged from the data. Research findings have been integrated and presented as thick descriptions complemented with verbatim quotations in this paper.

## 3. Results

### *Lagas* concentrate animals and result in the comingling of sick and healthy herds

*Laga* is one of the common pool resources within the drylands as exemplified by our study sites. Therefore, access and use of *laga* remain collective and communal. Their situation on routes to grazing lands, also, indirectly increases their usage by a variety of pastoralists. From a sociocultural lens, livestock watering assumes a collective shape and social behaviour patterned to benefit various households’ access to water a limited yet important common good. We observed a wide range of livestock, by species and origin, freely mingle as they await their drinking turns (see Plates 1& 2).

“…*multiple watering points dry up over the drought seasons, we all walk our herds to this point [laga] and water them [livestock] from the shared shallow wells” (informal interview with herdsman, Laisamis)*.

**Plate 1:**
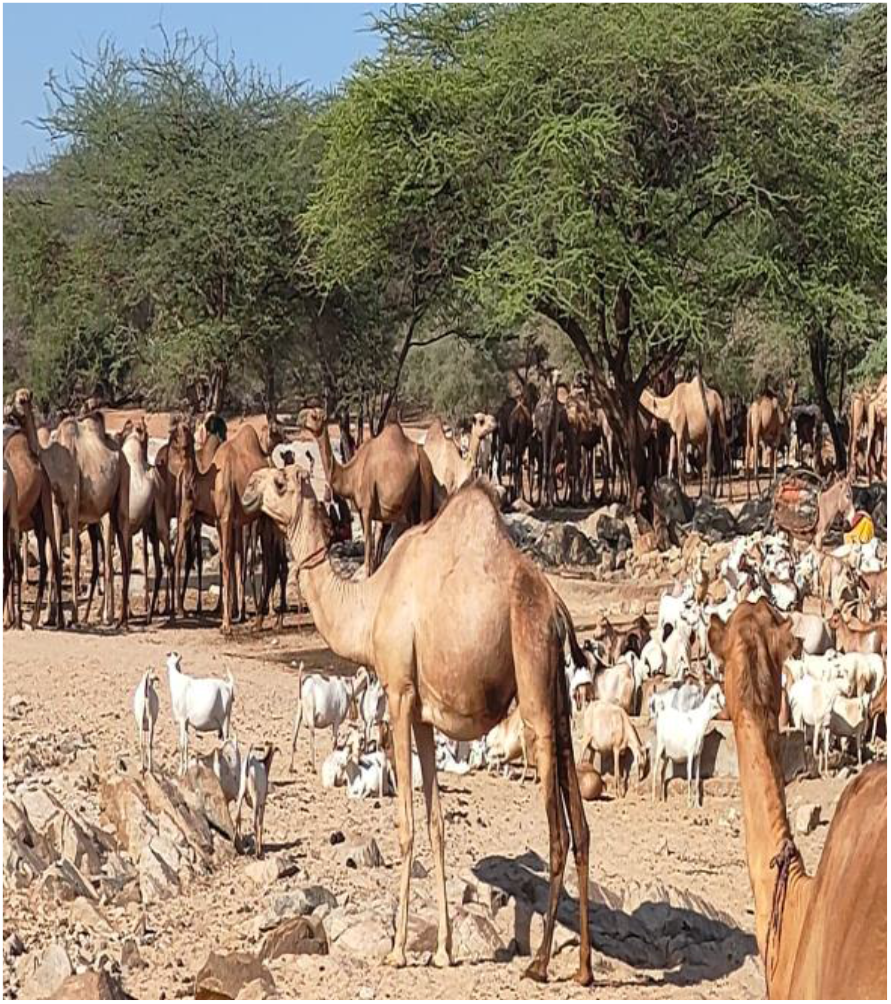
Livestock awaiting drinking turns in Laisamis

From a disease risk perspective, the relatively confined livestock movements and proximity increase the chances of mating as well as licking of vaginal discharges from potentially infected ones, which, in turn, increases the brucellosis transmission risks.

*“Many families depend on this laga, they walk animals for over 30 km from grazing fields…the water supply here is dependable, all types of animals lie on the laga awaiting their turns, even wildlife” (informal interview with herdsman, Laisamis)*

Additionally, we observed no separation of the sick herds from the healthy ones. The practice of non-isolation may be influenced by the pastoralists’ perceived low risk of brucellosis including its potential spread to other healthy herds. The latter translates into limited knowledge of the disease (brucellosis) signs and symptoms among community members. It might also be the fact that pastoralists know the symptoms; however, the need to see the livestock rehydrate and hopefully recuperate takes priority over isolation. Further, this non-isolation must be seen through the lens of *laga* being the only source of water within the locality for livestock, spatially displaced from the homesteads, hence, the need to have all livestock driven to the common resource.

**“***These animals belong to distinct families but they always move and get watered as a pool. There are no trained officers to monitor cases worth quarantining as they do in the markets… even the livestock owners will still push their sick herds till they cannot move anymore*.*” (Community Disease Reporter (CDR), Kajiado)*.

### Water troughs may facilitate transmission

Sharing of animal watering troughs (Plate 3) is common in low-resource settings. Troughs are composite construction designs of shallow wells, and where absent, animals often draw water from open surfaces on the *laga*. Trough sharing builds bonds since water is often dug into the *laga* surface and requires lots of manpower to extract; hence, materials and labour remain shared. Whereas the practice has great cultural and justifications, it poses the danger of cross-infections among livestock. There is a high risk that bacteria-infected saliva and nasal discharges end up being deposited in water and ingested by healthy herds predisposing the latter.

**Plate 2:**
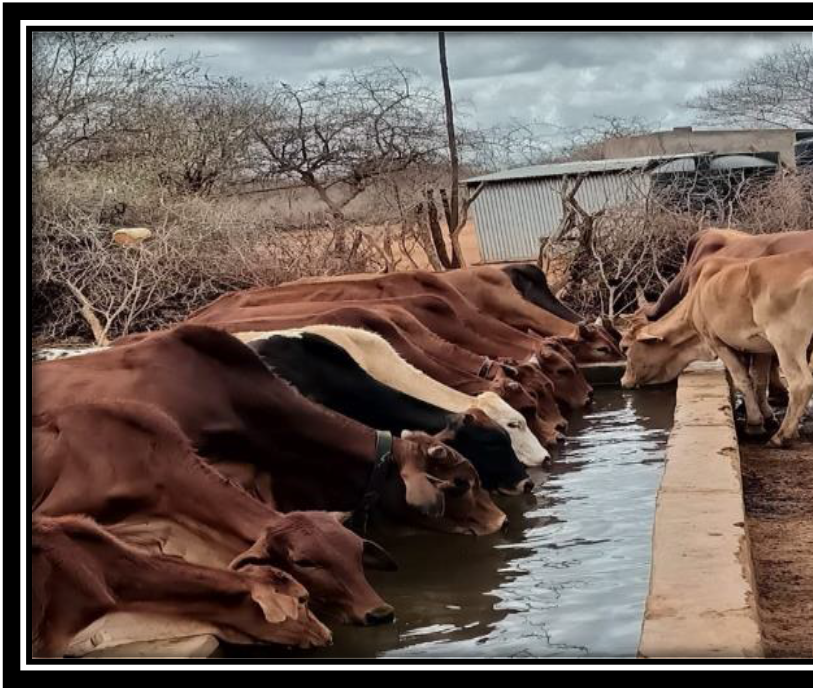
Cattle watering from a common trough in Mailua-Kajiado

*“Animals are watered at the same time, you need your herd to fill up and go back to pasture grounds. It becomes a problem if any of them is infected and leaves those bacterial droplets in water*.*”* (*KII, Laisamis)*.

We observed neither lay nor conventional mechanisms for disinfecting the watering troughs between herd transitions or before the first stock of herds that arrived at the *Laga*. Given the predisposition for wildlife use and depositions of *Brucella* from preceding herds, we hypothesise that these shared drinking troughs collectively serve the points of zoonotic disease transmission.

“*It is a case of cross- and re-infection, nobody inspects and advises pastoralists on safe handling of watering troughs. Remember, the same water will be drunk by wildlife who might also be infected, so we need to think of a way to manage bacteria by way of disinfection*.*” (interview with CDR Laisamis)*.

The above situation may create a continuous cycle of infection among various livestock groups including the wildlife using the same watering points. Indeed, the use of the troughs by wildlife is often overlooked and to an extent normalised as mutual co-existence default, yet, it forms such an important node in Brucella transmission to the domesticated herds.

**“**At the *laga, during the season, some wild animals like Zebra drink from the same troughs as domestic animals heightening the risks of cross-contamination” (Male IDI, Laisamis)*

There is a lay belief that livestock that normally graze together pose limited chances of infecting one another even if they drink from a common pool. It might loosely translate to the perception that disease threats will always emerge from herds external to the common grazing and watering fields. This points to limited knowledge among occupationally at-risk populations. While they (herders) are in close contact with livestock including watering, their practice of mingling, and pooled watering of livestock as a unit makes them a weak link in brucellosis prevention and invokes the need for exposure behaviour training by veterinary and human health divisions.

“*If it were such a driver of disease contraction, they would have all been infected, this is how we have always watered our livestock, they graze and drink together, it is the same herd*.*” [Informal interviews with herdsman, Kajiado]*.

### *Lagas* exacerbate contact between different animal species

It is common to spot different animal species, cattle, camels, goats and sheep, from different satellite grazing fields driven to the same watering point. Additionally, the wildlife from the nearby rangelands and conservancies drink water from the same watering points. Once at the laga, a variety of small and large ruminants freely interact and sometimes graze on the grass-patched areas of the l*aga* (Plate 3).

**Plate 3:**
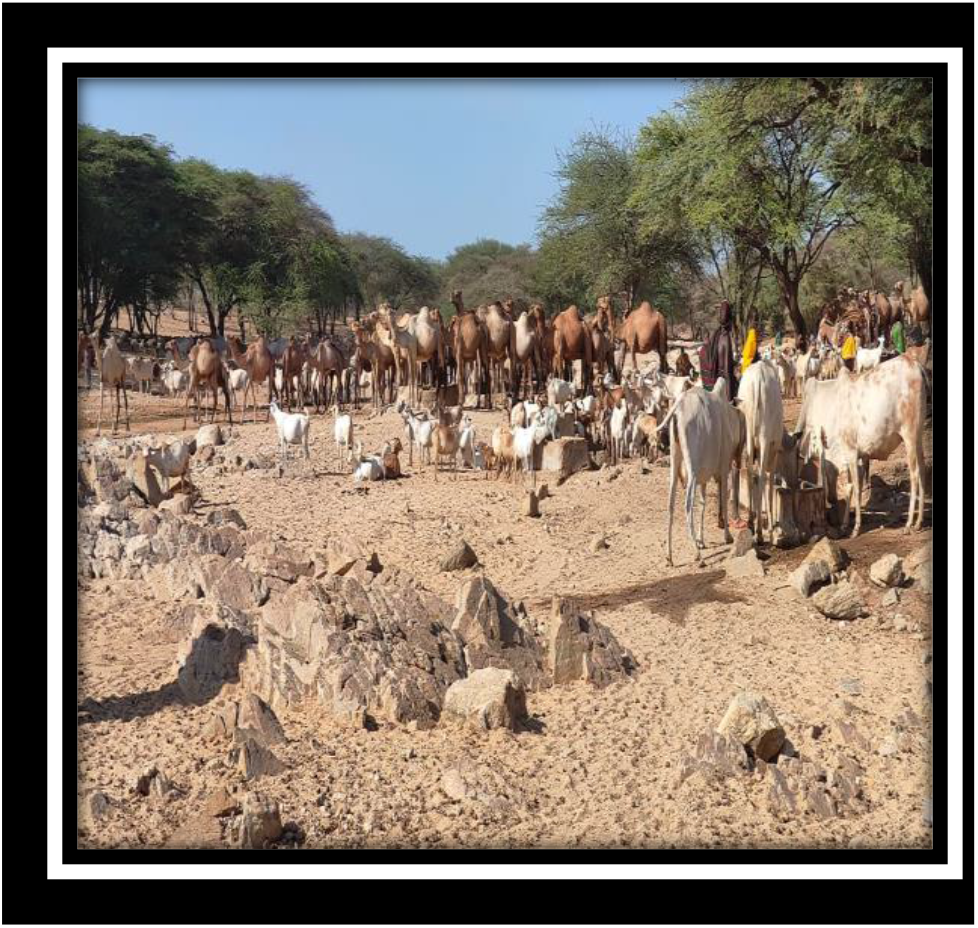
Multi-species mixing at Laisamis Laga

The risks here are four-fold. First, there is the risk of transmission on the route to watering points especially if an infection is picked from the grazing fields. Second, there is the risk of contracting brucellosis as animals freely mix as they await their turns to drink. Third, cross-species sharing of watering points increases the Brucella transmission risks with small ruminants always seen as major hosts. Fourth,

*“Goats and sheep often host a lot of bacteria. In the laga, you will see them freely share the water with other species, which translates to high-level cross infections*.*” (KII, Laisamis)*.

This mixing occurring at the inter-species level, is oblivious of the increased transmission risks across the herds. The presence of different animal species, including domestic and wild animals at the *laga* increases the potential for disease transmission. Some kind of mutual co-existence occurs among humans, domesticated livestock and wildlife in terms of shared waterpoints. From a conservation lens, it is the sound thing to do, to allow the wildlife to drink from the same sources during the night, from an epidemiological prism, the risk of Brucella spread from unvaccinated and unmonitored wildlife is tenfold. Consequently, we note the health risks that this otherwise conservation utilitarian value poses to humans and animals at the laga.

### Feeding dogs on abortuses and placental materials may contribute to disease spread

Proper and safe disposal of parturition materials and abortuses is key to the prevention of the spread of brucellosis given the high concentration of *Brucella* in the reproductive systems. Community narratives point to the normalisation of feeding dogs on what’s deemed as ‘waste’ ‘inedible aborted and placental materials. The feeding of dogs with such materials not only risks infecting the dogs themselves but also increases the likelihood of transmission to humans or other animals through close contact or exposure to contaminated materials. Despite these risks,,to many pastoralists, this activity seems harmless to the dogs and provides a more meaningful disposal of animal products that humans cannot consume.

> “*All that waste together with the placenta is given to the dogs*.*” (Female, IDI Kajiado)*.
>
> *“If you go to any community, they had ways of doing things out of ignorance. For example, if it is an abortus, it’ll be given to a dog because, to them, it’s meat*.*” (KII, Laisamis*).

It is also deemed beneficial in resource-scarce settings where the dogs normally scavenge for their food, hence, maximization on feeds even where it presents inherent risks always goes on unabated. The practice might also follow some dietary prohibitions. In Marsabit, for instance, the Rendile do not consume some organs like lungs, spleen and glands, which often end up being fed to the dogs making the latter susceptible to Brucella.

“*I let them-placenta-fall then roll them on a stick and given them [placenta materials] to the dogs” (Male IDI, Kajiado)*.

Additionally, we noticed weaknesses in placental disposal practices for those who cared to bury the same upon parturition. This involved the use of shallow disposal pits dug by the *laga* where the placenta is buried. Additionally, the disposal did not involve any form of burning to get rid of the bacteria. The near-ground surface burials are also seen to be a risk factor among scavenging dogs while at the same time contaminating the *laga* ecosystem with *Brucellae* in case of infected placental materials.

> *Yes, even that animal which has been aborted …they don’t dig like a grave, just a small thing which the dog will come and just scratch. (KII, Marsabit)*.

### Herd to human risk behaviours and practices

#### Abortus handling at the laga risk Brucella spread to humans

Improper management of aborted material from infected animals plays a pivotal role in the transmission of brucellosis. Handling aborted material without protective equipment poses a significant risk of transmitting brucellosis and other zoonoses since the bacteria are present in the placental tissues, vaginal discharges, and foetal fluids in high concentrations. The risks of *Brucella* transmission are heightened if there are open/cut wounds on their hands. Most animals give birth within the grazing fields and on the *laga*. Herdsmen, with experience delivering animals frequently volunteer their services to assist with parturition, potentially increasing the risk of transmission between herds.

*“There was a time I found my neighbour’s goats struggling to deliver, I pulled out the young one and carried it home (neighbour’s)…it is a common thing to do… just bare hands*.*” (informal interview with herdsman, Kajiado)*.

**Plate 4:**
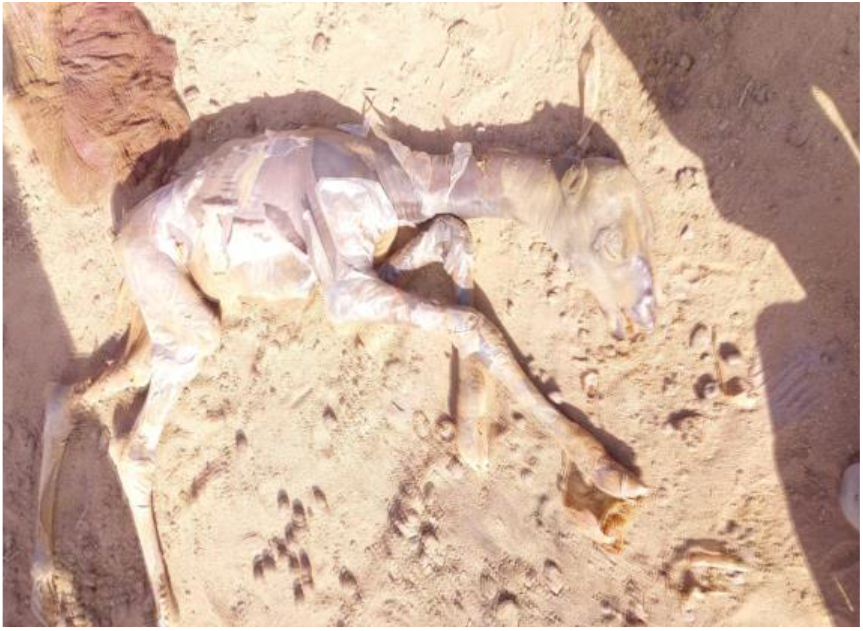
Camel abortus lie by Laisamis laga

Because personal protective devices and disinfection are generally unavailable, this practice can facilitate the transmission of the bacteria to humans. Handling infected abortus with bare hands can increase the likelihood of bacteria entering the body, especially if one inadvertently touches the face.

> *“It is always bare hands. Delivering is done with bare hands and they can even go to the extent of kissing and aspirating a calf if they feel that it has amniotic fluid within its lungs. They will directly suck the way people suck the blocked nostrils of small children*.*”* (*KII, Kajiado*). “*Sometimes you find a cow giving birth…and the birth process won’t release the calf, there is an elderly man who comes to deliver the cow and rectifies the situation…” (IDI, Kajiado)*.

#### Using urine as an antiseptic may provide avenues for infection

Urine has multiple uses within pastoral settings especially that of cattle and camels. The use of camel urine as an “antiseptic” for treating open wounds is rooted in old age tradition among the Rendile community. The urine is believed to contain substances with healing benefits on wounds due to its burning sensation. Across the two sites, animal urine is normalised for cleaning the udders before milking and at times washing off the surfaces of milking devices.

> “*For morans (young male warriors in maasai and samburu community), the urine, especially for camels is medicinal, when they get scratches on their hands, they tickle the camels to pee on the infected surface. The wound eventually dries up…we do not understand what happens but that is common over here” (Male CDR in Laisamis*).
>
> *“Some use the urine of the cow to clean the hands and proceed to milk the animal*.*” (Male IDI, Kajiado)*.

There are, however, significant health risks associated with it the use of urine on wounds particularly concerning the spread of *Brucella* and other zoonoses-causing pathogens due to the high concentration of bacteria present in the urine of infected animals (Salisu et al., 2018). Its application as medicine introduces a direct route for the transmission of *Brucellae*. When applied to open wounds, the bacteria can easily enter the bloodstream, causing infection and leading to the onset of brucellosis in humans. Moreover, the nature of wounds, being open portals to the internal system, increases the vulnerability to bacterial infiltration, further facilitating the spread of the disease between herds and humans.

#### Dietary habits: raw milk and blood consumption

Some milking and consumption of raw milk goes on at the watering points. We observed cases where women who came to replenish the herdsmen’s food supplies engaged in milking goats or collected milk from the camels to consume and also ferry back to other household members left behind. This practice highlights cross-herd consumption, as milk is sourced from various animals belonging to different herds. The incidence of instant milk consumption was also evident among the herdsmen despite the risk factors such practice portends.

“*The camel milk is rich [nutritious] and pure, it keeps us healthy…you drink it hot from the camel because if you boil, it will not taste the same” (Informal interview with herdsman, Laisamis)*.

The milking preparation process seems to oscillate between risk-reduction and risk-retention practices embedded in cultural prisms of what works in eliminating bacteria from hand surfaces.

> *“These days we use water and Omo [detergent] to wash our hands but earlier we used to use the dry cow dung to clean our hands*.*” (Male IDI, Kajiado*).

The question of sanitising the hand also assumes gender and species dimensions. It emerged in the study small animals like goats are freely, milked without cleaning hands, unlike the big ruminants, cows and camels, milked by men. Even among the latter species, it is the cleaning agent, urine, that still poses the risk of *Brucella* transmission. Whereas hand hygiene practices are largely done to reduce pathogen transmission to animals, the reverse risk of Brucellosis for humans via the use of urine is hardly thought through, a grey area in public health intervention that requires re-thinking within water-scarce environments.

> “*It is women who milk the goats, they hardly wash their hands, it is men who normally wash their hands with urine from the cattle or camels before milking (Female IDI, Laisamis)*.

A preventive measure across the two sites is the use of ash and herbal products from specific tree species to disinfect the milking guards which act as temporary storage facilities.

> *“…the milking guard is normally prepared[cleaned]using some tree leaves and burnt charcoal to ensure the milk does not go bad [not contaminated] …,” (Male IDI, Marsabit*).

The milk from the *laga* does pose a threat of spatial infection beyond the site. It is often taken back to the family to supplement their dietary needs. However, we note that such milk might be mixed, drawn from different cows including the sick animals, which is another stream of infection root.

> *“The milk is mixed so you don’t know which animal among them is sick it’s only in camels that they milk… sometimes when the milk is from a camel and it’s a very small amount it’s mixed with another animal’s like a goat if they belong to the same person’’ (KII, Marsabit*).

### *Laga* fomites

#### Contaminated livestock excreta

Animal excreta harbourings a wide range of bacterial pathogens are constantly deposited on the *laga* surface (Plate 5) where we see relatively dry camel manure in Laisamis-Marsabit and water channels (Plate 6) where we see a watering point polluted by animal manure in Mailua-Kajiado. Indeed, defecation on a watercourse may have spatial effects on disease spread especially where such deposits are swept away by water currents. Both situations, Plates 1 and 2 present significant risks for the transmission and spread of brucellosis. Bacteria can survive for extended periods in manure, allowing it to contaminate soil and water sources, thereby posing a considerable threat to animal, human and environmental health.

“*There is no management of animal manure deposits on the laga, perhaps, people don’t see these as potentially hosting bacteria, yet, they form a good environment for Brucella to survive till it finds a new host” (KII, Marsabit)*.

Infected animals usually excrete waste on the *laga* and exposure to these contaminated excreta, either through direct contact or grazing on grass on the *laga*, can lead to healthy animals contracting the bacteria.

*“It is not how healthy your herd is, the environment from where they are watered can be contaminated like with untreated animal manure which poses the transmission danger*.*” (KII, Kajiado)*.

#### Contamination of *laga* sand and stones with parturition materials

Small animals, like goats, sometimes give birth at the watering site which may contribute to the environmental contamination (Plate 7), with brucellosis-causing bacteria. As noticed during the handling and disposal process where abortuses are not disposed of properly, *Brucella* species can contaminate the surrounding environment by indirect transmission through soil, water, or feed contamination as seen with the poor waste management practices in the areas. Other animals are exposed to the disease when they lick the surfaces where an animal has given birth. Sand contaminated with bodily fluids from infected animals, such as afterbirth, urine, or faeces, becomes a breeding ground for the Brucella bacteria. When livestock come into contact with this sand, they can easily contract the bacteria.

**Plate 5:**
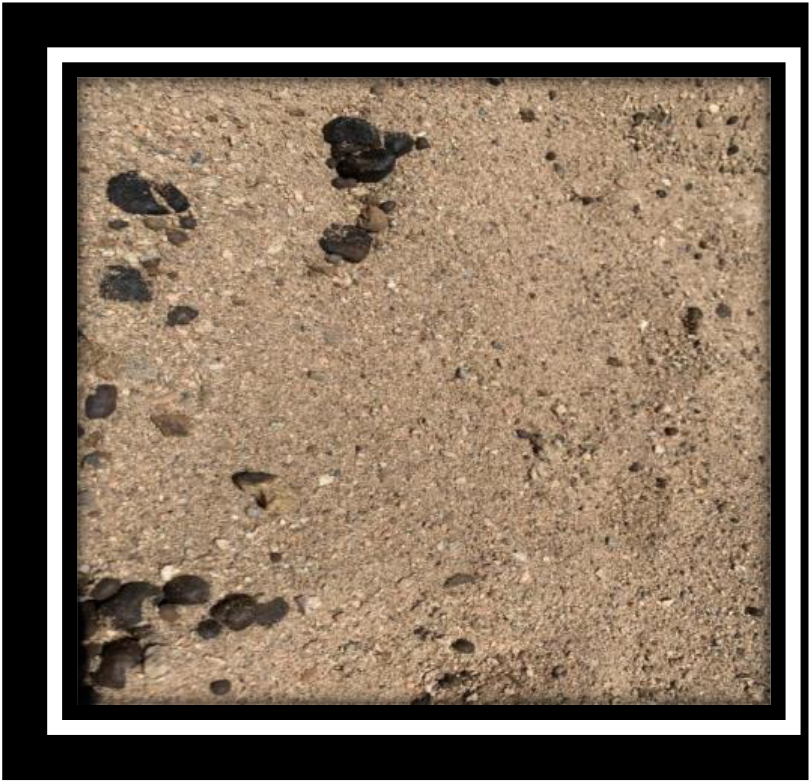
Camel droppings in Laisamis Laga

**Plate 6:**
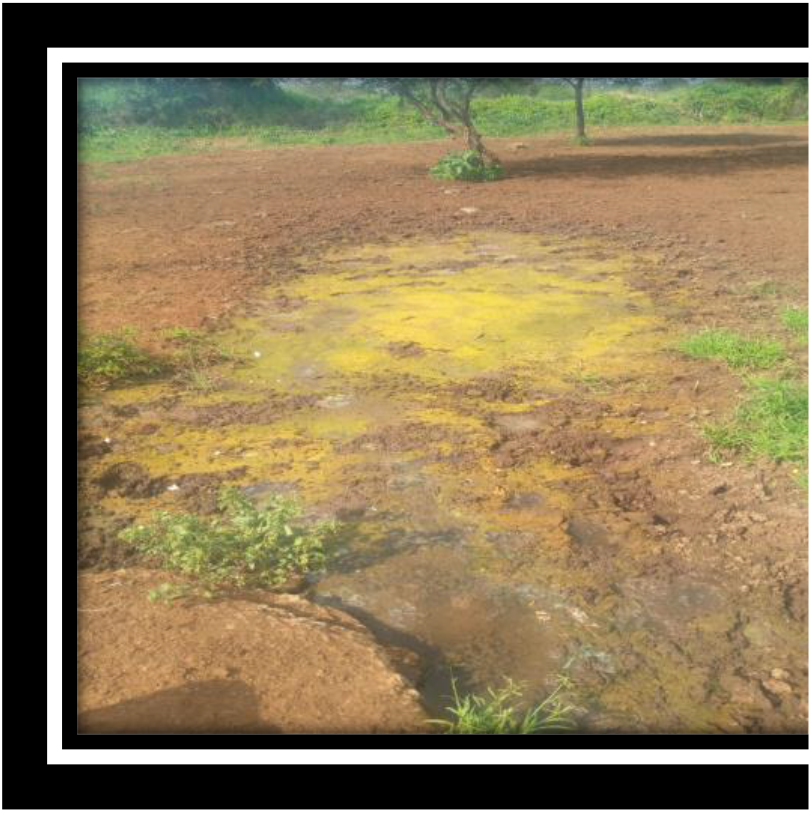
Manure contaminated water in Kajiado

**Plate 7:**
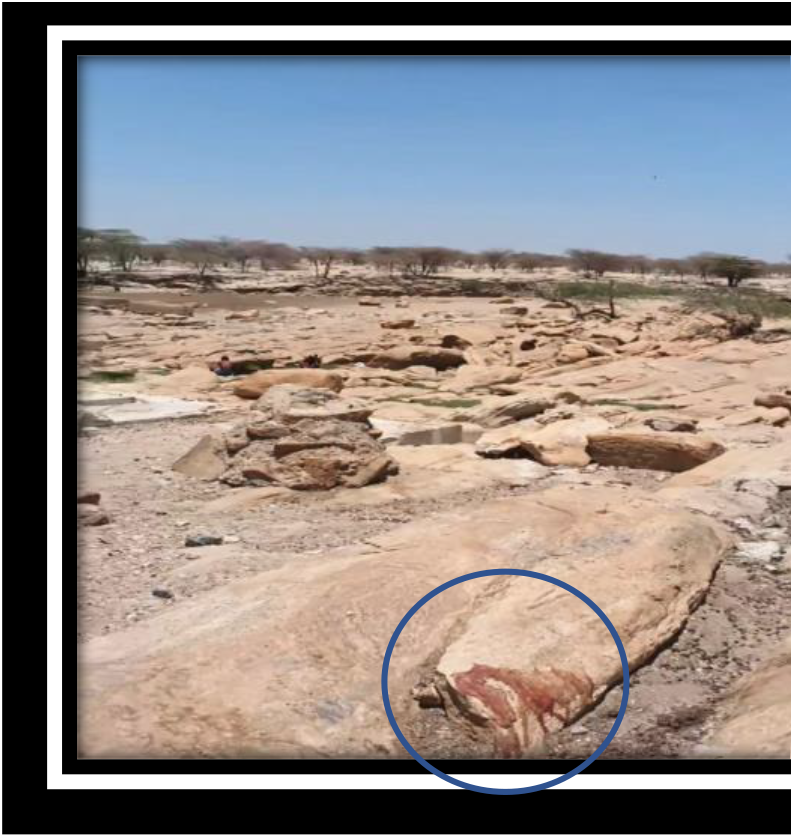
Parturition materials of a goat in Laisamis *Laga*-Marsabit

“*There are so many animals giving birth and contaminating the rocks and the nearby sand. The materials are swept back into the laga or other animals simply lick them up and they pose high risks of brucellosis transmission”* ***(KII, Marsabit)***.

We learnt of the community’s preference to clean fresh hides from the slaughter slabs by the *laga*. It follows that the hot and wet sand offers a better abrasion against any flesh stuck on the hides. This practice while cheap and common, risks contaminating the *laga* sand with bacteria washed off the animal skins. It further aggravates the cycle of infection as contaminated sand acts as a reservoir for the bacteria because animals that come into contact with the contaminated sand can become hosts of the bacteria with the potential for spatial spread.

**“***At the peak of slaughtering, people come over here (laga) to clean the hides before they can go and dry them at home, it contaminates both the water and the sand, and we end up with bacteria transferred to our livestock” (interview* with CDR, Laisamis).

#### Contaminated surface water

Contaminated surface water (Plate 8), can serve as a haven for the transmission and spread of Brucella because the bacteria can survive for extended periods in water, especially in stagnant surfaces as seen on the *laga*. As livestock infected with brucellosis excrete bodily fluids like urine, faeces, or placental tissues into water bodies, these contaminated fluids can introduce bacteria into the water, creating a potential hazard for human exposure since the water is used for drinking and domestic purposes. The same water can be hazardous to other livestock that drink from these sources.

“*the contamination is very high especially where animals give birth and discharge a lot of bodily secretions, we may need to expose a lot of those materials from the laga to lab test and establish if Brucella is among the shed organisms “(KII, Marsabit)*.

**Plate 8:**
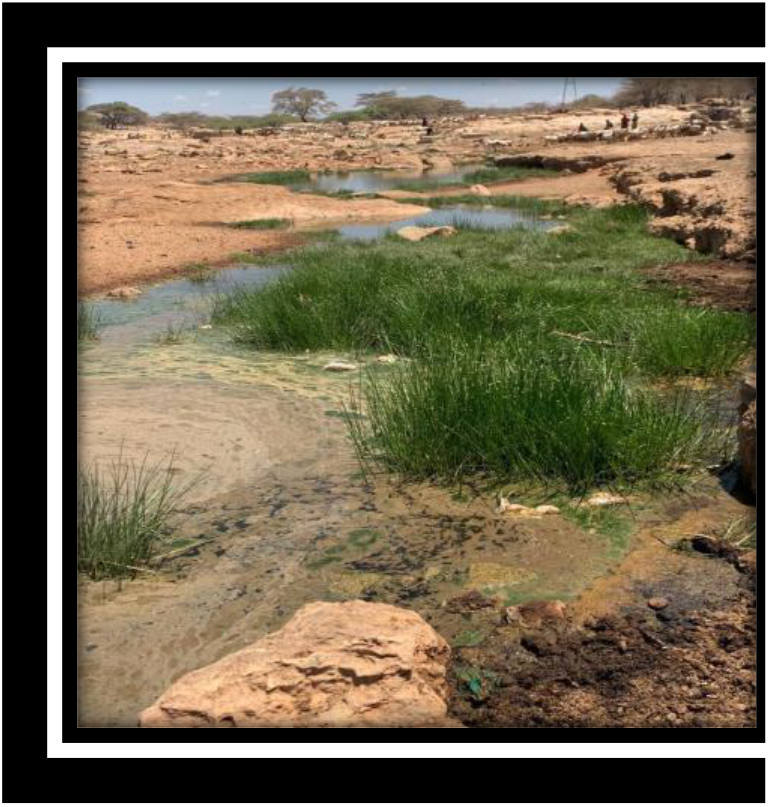
contaminated water recharging laga ecosystem

#### Infected aborted foetuses on the *laga*

Infected aborted foetuses discarded on the laga pose a significant transmission risk since brucella can survive on the surface for extended periods. As scavengers and other pet animals like dogs consume these remains, they become potential carriers, perpetuating the cycle of infection and increasing the likelihood of inter-herd and intra-herd brucellosis transmission. In the ecosystem of the laga, scavengers, dogs and wildlife are drawn to carrion, including aborted foetuses, as a food source. This scavenging behaviour poses a substantial risk of spreading brucellosis. As scavengers feed on infected tissues, they can become carriers of the bacteria, allowing the pathogen to persist in the environment hence increasing the risk of exposure for healthy animals.

## Discussion

We aimed to depart from the binary clinal debate inspired by pathogen theory to a socio-cultural prism on human-animal health concerning the prevention and control of brucellosis, particularly, considering the pathways of *Brucella* species transmission. This departure shifts the focus from viewing infectious diseases solely as biological phenomena to understanding the complex relationship of social, cultural, and environmental factors in disease transmission dynamics. We bring on board two more loci, the environment and local-situated reality, informing risky practices in brucellosis transmission (Godfroid et al., 2011; Adisasmito et al., 2022). Our approach further affords assessment of risk behaviors *in situ* to elevate localized perceptions in public health messaging. Consequently, the study argues that optimal preventive measures and health for humans, livestock and the environment require insights from social and bio-clinical evidence (Nyamongo, 2002). The study achieves the former via the co-production of knowledge (Bergeron et al., 2014) through in-depth multi-ethnographic methods deployed in the current study. The study aims to inspire the latter through further clinical analysis of risk factors present within the larger *laga* ecosystem. Such fusion of evidence strengthens the response to the endemicity of brucellosis in Africa since it is grounded on multidimensional knowledge.

The level of awareness among pastoralists on brucellosis cannot be underestimated. However, their understanding of certain aspects, such as the contagious nature of the disease, remains limited despite their overall knowledge (Helman, 2007). This limited understanding contributes to the persistence of risky practices at the *laga* and the continuation of less precautionary behaviors observed among pastoralist communities (Solera et al., 1998; Saleem et al., 2010). For instance, limited knowledge on the longevity of the bacteria on the water surface, excreta, parturition materials and other contaminated solid materials continue the continuum of risks for *Brucella* species transmission within herds and to humans. It is also evident that the residents have little knowledge of the risks of intra and inter-herd transmission of diseases except via drinking of raw milk and consumption of infected meat. Even where such fears of transmission exist, the ecological settings defined by limited watering points except for dependable *laga*, especially during extended dry seasons naturally gravitate toward close interaction within herds and across species pointing to an environmental tributary of *Brucella* transmission source. Therefore, continued climate variability will intensify such interactions. Further, the study acknowledges daily livestock movement and communal watering behavior, which essentially are adaptive within drylands, and embedded in complex community systems of dependence on common pool resources exemplified by the *iaga* (Manika and Gregory-Smith, 2017), all of which increase the risk of *Brucella* transmission.

Perceived risk is also a factor in the potential human contraction of brucellosis at the *laga* especially in the etiology (Obonyo and Gufu, 2015) and transmission pathways. Whereas clinical studies point to infections in humans via abraded skin (CFSPH, 2018), we observed the constant use of urine as an antiseptic against open wounds. While the practice can be hailed as part of ethnomedicine, it has to be cautioned for its transmission potential. The presence of Brucella bacteria in camel urine, as indicated by previous studies (Salisu et al., 2018; Gutema and Tesfaye, 2019), emphasizes the potential role of camel urine as a source of Brucellosis transmission. Further, the study observed the potential risks of *Brucella* transmission through drinking from the same animal troughs, and bare-hand parturition assistance of animals including peeling off the membrane of the goat-kids. The latter practice represents high risk given Brucella is concentrated within the reproductive systems of animals and shed off in high volumes during parturition (Eko et al., 2022) The study noted that herders who are the primary caregivers for animals, were more likely to perform the above tasks and engage in risky behaviors including assistance in delivering livestock increasing their occupational risks (Osoro et al., 2015) within the *laga* ecosystem.

Three features of the *laga* increase comingling of animals and may increase the risk of brucellosis transmission. First, water at the *laga* constitutes a common resource pool that holds the lifeline for all living organisms in the dry rangelands. Its access and consumption remain universal and predicated on survival despite the underlying brucellosis transmission risks. Second, is the community ownership of the wells at the *laga* and the pooling of labor necessary to get the animals their fill. The morans, young herders, are tasked with filling up the watering troughs, while women milk animals for home consumption and replenish food supplies for the morans returning to distant grazing lands; third, is the systemic failure of the veterinary and public health sectors to erect screening and sensitization booths around the watering points where overlayered risks of *Brucella* transmission peak. Whereas the study observed that herds take turns drinking, it does not limit pre-drinking interactions or eliminate transmission risks arising from the contaminated fomites.

Mixing of herds also promotes inter-specific interactions within the *laga*. We observed multiple cases of small ruminants naturally blending with the larger varieties. Despite bio-clinical evidence (CFSPH, 2018) pointing to the risks of cross-species infection under such arrangements, the practice of mixed farming and watering is socio-economically and ecologically rationalized. First, these small ruminants provide essential macro and micro-nutrients crucial for the health of children, infants, and women of childbearing age (Alonso et al., 2019; McKune et al., 2022), particularly in dry rangelands where such nutrients are vital for survival. Therefore, their contribution cannot be overlooked. Second, small ruminants serve valuable uses like cultural and religious meanings, social status, and asset accumulation (Alders et al., 2021; Schneider and Tarawali, 2021). Finally, it is these species that normally survive long dry seasons better than bigger ruminants and remain in fallback positions for families. Thus, we note that the risk of cross-species transmission cannot be evaluated over these socio-economic and ecological values and/or cultural significance of mixing. What needs deepening is the safe animal husbandry practices involving the isolation of sick herds and vaccination to sustain healthy production and eradicate brucellosis risks.

The practice of milking, drinking and storage for subsequent consumption also occurs at the laga. While not unusual among pastoral communities, the laga presents a unique restocking point for the families left behind, especially the children and women, while at the same time, the herdsmen also feed before starting their return journeys to the grazing lands. It is the unprocessed milk consumption as dietary behavior and storage practices that we perceive as presenting potential brucella transmission pathways. Community-level cultural and nutritional justifications have been advanced to back up this practice. Livestock milk is preferred for its taste (Njenga, 2020) and is believed to provide hydration and boost immunity in young children (Onono et al., 2019). Additionally, it is not associated with any reported casualties (Mburu et al., 2021). Given the ecological settings, often dry and largely food-deprived, animal-source foods such as milk play vital survival roles. What is less discussed is the risks associated with this raw consumption practice and the potential way of transitioning the behavior to cut down on potential brucellosis transmission. This practice seems to cut across generations and social classes but peaks much with the elderly. Further, there is a lull in scholarship on the effects of herbal products used as milk preservatives on the bacteria in the case of storage.

In the study, we noted continual contamination of the *laga’s* physical body-the water, sand, grass, watering troughs, and rocks-all of which affect the environment and that of humans and animals dependent on it. Importantly, we noted that bacteria are potentially passed on to the body via bodily fluid discharges from livestock, parturition materials including placentae, animal droppings (manure), and human footprints during water abstraction and bathing. Essentially, the anthropogenic and livestock activities not only put pressure on a fragile yet important ecosystem for the pastoralists but also create favorable conditions for the re-emergence of brucellosis. It is also worth noting that during extended dry seasons, over which the current study was carried, the wind blows across the *laga* exacerbating the risk of bacteria inhalation.

### Limitations of the study

Our study draws from two pastoral communities in Kenya, namely; the Maasai and Rendile, which methodologically, might not capture the diversity of pastoral communities, and therefore, limit the transferability of the findings. However, the near-universal cultural practices of pastoralists and their tendency to occupy dry rangelands (USAID, 2020) gives the impetus to believe these transmission risks are universal and applicable across space and cultures in low- and middle-income countries where pastoralism is practiced.

## Conclusion

The study appreciates the centrality of the *laga* ecosystem in sustaining the lives of humans, animals and the environment. We reiterate the fact that it is a window for examining multispecies entanglements under One Health and re-imaging brucellosis preventive behaviours and control strategies within the drylands. This study indicates the need to have clinical samples at multiple sites of the *laga* to check for Brucella including the spatial distribution along the *laga* channel. It also primes for the inclusion of local perspectives in public health and animal health interventions given the emic, insider, values in rationalizing laga-based practices. Reframing the narratives of brucellosis risk communication must be shaped by these cultural and ecological realities in the dry rangelands. The study proposes the use of the fear appeals framework in examining the brucellosis threats within the *laga* ecosystem as part of informing the behavior change and risk communication under One Health.

From a methodological standpoint, we emphasize that our non-biomedical perspective gives room to interrogate behaviors *in situ* and put into focus the localized understanding of *Brucella* transmission risks. This is significant in bringing out the more often marginalized voices in preventive public health policy framing and preventive behavior messaging. Further, we have demonstrated that *laga* risk factors have a potential for transmission along the water channel, therefore, we advocate for spatial testing for Brucella presence at multiple sites along the *laga* channel.

## Data Availability

The data used in this work is de-identified and available at WSU-Global Health Kenya upon formal request.

## Acknowledgement

The authors would like to extend their gratitude to the Marsabit County government, the County Government of Kajiado and study staff based at Washington State University Global Health, Kenya. The research team appreciates the support from human and animal health workers based in Kajiado and Marsabit Counties who gave unwavering support and link to the communities. The research has immensely benefited from the funding by the US Defense Threat Reduction Agency award # HDTRA12110041.

## Data availability

The data used in this work is de-identified and available at WSU upon formal request.

## References

Abubakar, M., Mansoor, M., & Arshed, M. J. (2012). Bovine Brucellosis: Old and New Concepts with Pakistan Perspective. Pakistan Veterinary Journal, 32(2).

Adisasmito WB, Almuhairi S, Behravesh CB, Bilivogui P, Bukachi SA, et al. (2022) One Health: A new definition for a sustainable and healthy future. PLoS Pathog 18(6): e1010537. 10.1371/journal.ppat.1010537.

Alonso, S., P. Dominguez-Salas, and D. Grace (2019). The role of livestock products for nutrition in the 1000 days of life. Anim. Front.9(4):24-31. Doi:10.1093/af/vfz033.

Anderson J, Jones K. The difference that place makes to methodology: uncovering the ‘lived space’ of young people’s spatial practices. Child Geography, 2009;7(3):291–303.

Bergeron J, Paquette S, Poullaouec-Gonidec P.(2014) Uncovering landscape values and micro-geographies of meanings with the go-along method. Landsc Urban Plan. 2014;122:108–21.

Centers for Disease Control and Prevention (2019). Brucellosis. National Center for Emerging and Zoonotic Infectious Diseases (NCEZID), Division of High-Consequence Pathogens and Pathology (DHCPP). Available at https://www.cdc.gov/brucellosis/transmission/index.html Accessed on 24-01-2024.

CFSPH (2018). Brucellosis: Brucella Suis. Available at https://www.cfsph.iastate.edu. Accessed on 13-12-2023.

Diseases and Exposure Assessment to Brucellosis within rural and peri-urban areas in Kajiado, Kenya. F1000Research, 8, 1916.

Djangwani J., Abong’ G.O., Njue L.G., Kaindi D.W.M. Sero-prevalence and risk factors of Brucella presence in farm bulk milk from open and zero grazing cattle production systems in Rwanda. Vet. Med. Sci. 2021;7:1656–1670. 10.1002/vms3.562.

Djangwani, J., Ooko Abong’, G., Gicuku Njue, L., & Kaindi, D. W. M. (2021). Brucellosis: Prevalence with reference to East African Community countries - A rapid review. Veterinary medicine and science, 7(3), 851–867. 10.1002/vms3.425.

Eko, S. M., Esemu, S. N., Nota, A. D., & Ndip, L. M. (2022). A review on brucellosis in Cameroon: diagnostic approaches, epidemiology and risk factors for infection. Advances in Microbiology, 12(7), 415–442.

Electoral, I. (2012). Preliminary report on the first review relating to the delimitation of boundaries of constituencies and wards. Nairobi, Kenya.

El-Sayed, A., & Awad, W. (2018). Brucellosis: Evolution and expected comeback. International journal of veterinary science and medicine, 6, S31–S35.

Food and Drug Administration. (2012). Bad bug book: Handbook of foodborne pathogenic microorganisms and natural toxins. In U.S. Department of Health and Human Services (2nd ed.). 10.1016/S1872-2040(10)60451-3.

Franc, K. A., Krecek, R. C., Häsler, B. N., & Arenas-Gamboa, A. M. (2018). Brucellosis remains a neglected disease in the developing world: a call for interdisciplinary action. BMC public health, 18, 1–9.

Fratkin, E., Roth, A. E., & Nathan, M. A. (2004). Pastoral Sedentarization And Its Effects On Children’s Diet, Health And Growth Among Rendile Of Northern Kenya. Human Ecology-, 531–559.

Godfroid, J., Debolle, X., Roop, R. M., O’callaghan, D., Tsolis, R. M., Baldwin, C. J., Santos, R. L., Mcgiven, J. A., Olsen, S. C., Nymo, I. H., Larsen, A., Al dahouk, S., & Letesson, J. J. (2014). The quest for a true One Health perspective of Brucellosis. OIE Revue Scientifique et Technique, 33(2), 521–538. 10.20506/rst.33.2.2290.

Godfroid, J., Scholz, H. C., Barbier, T., Nicolas, C., Wattiau, P., Fretin, D., … & Letesson, J. J. (2011). Brucellosis at the animal/ecosystem/human interface at the beginning of the 21st century. Preventive veterinary medicine, 102(2), 118–131.

Gutema, F., & Tesfaye, J. (2019). Review on camel brucellosis: public health importance and status in Ethiopia. Academic Research Journal of Agricultural Science and Research, 7(7), 513–529.

Helman, C. (2007). Culture, health and illness. CRC press.

Lokamar, P. N., Kutwah, M. A., Munde, E. O., Oloo, D., Atieli, H, Gumo S, Akoko, J., and Ouma, C. (2022) Prevalence of Brucellosis in livestock keepers and domestic ruminants in Baringo County, Kenya. PLOS Glob Public Health 2(8): e0000682. 10.1371/journal.pgph.0000682.

Manika D., Gregory-Smith D. Health marketing communications: An integrated conceptual framework of key determinants of health behaviour across the stages of change. J. Mark. Commun. 2017; 23:22– 72. 10.1080/13527266.2014.946436.

Matle, I., Ledwaba, B., Madiba, K., Makhado, L., Jambwa, K., & Ntushelo, N. (2021). Characterisation of Brucella species and biovars in South Africa between 2008 and 2018 using laboratory diagnostic data. Veterinary Medicine and Science, 7(4), 1245–1253.

Mburu, C. M., Bukachi, S. A., H. Tokpa, K., Fokou, G., Shilabukha, K., Ezekiel, M., & AMP Kreppel, K. (2021). Lay attitudes and misconceptions and their implications for the control of brucellosis in an agro-pastoral community in Kilombero district, Tanzania. PLOS Neglected Tropical Diseases, 15(6), e0009500. 10.1371/journal.pntd.0009500

Meltzer, E., Sidi, Y., Smolen, G., Banai, M., Bardenstein, S., & Schwartz, E. (2010). Sexually transmitted brucellosis in humans. Clinical infectious diseases, 51(2), e12–e15.

Muturi, M., Bitek, A., Mwatondo, A. et al. Risk factors for human brucellosis among a pastoralist community in South-West Kenya, 2015. BMC Res Notes 11, 865 (2018). 10.1186/s13104-018-3961-x

Mwatondo A, Muturi M, Akoko J, Nyamota R, Nthiwa D, Maina J, Omolo J, Gichuhi S, Mureithi MW, Bett B. Seroprevalence and related risk factors of Brucella spp. in livestock and humans in Garbatula subcounty, Isiolo county, Kenya. PLoS Negl Trop Dis. 2023 Oct 16;17(10):e0011682. 10.1371/0011682

National Drought Management Authority (NDMA, 2023). 2023 Long Rains,Food and Nutrition Assessment Repoprt. Available at http://knowledgeweb.ndma.go.ke/ Accessed on 25/01/2024.

Njenga MK, Ogolla E, Thumbi SM, Ngere I, Omulo S, Muturi M, Marwanga D, Bitek A, Bett B, Widdowson MA, Munyua P, Osoro EM. Comparison of knowledge, attitude, and practices of animal and human brucellosis between nomadic pastoralists and non-pastoralists in Kenya. BMC Public Health. 2020 Feb 24;20(1):269. 10.1186/s12889-020-8362-0

Njeru, J., Wareth, G., Melzer, F., Henning, K., Pletz, M. W., Heller, M., and Neubauer, H. (2016). Systematic review of Brucellosis in Kenya: disease frequency in humans and animals and risk factors for human infection. BMC Public Health 16, 853. 10.1186/s12889-016-3532-9.

Nyamongo, I. K. (2002). Health care switching behaviour of malaria patients in a Kenyan rural community. Social science & medicine, 54(3), 377–386

Obonyo, M., & Gufu, W. B. (2015). Knowledge, attitude and practices towards brucellosis among pastoral community in Kenya, 2013. International Journal of Innovative Research and Development, 4(10), 375–84.

OIE. Brucellosis (B. abortus, B. melitensis and B. suis) (2016). https://www.oie.int/en/animal-health-in-the-world/animal-diseases/Brucellosis/.

Olsen SC, Palmer MV. Advancement of knowledge of Brucella over the past 50 years. Vet Pathol. 2014;51(6):1076–1089. 10.1177/0300985814540545

Onono, J., Mutua, P., Kitala, P., & AMP; Gathura P. (2019). Knowledge of Pastoralists on Livestock Diseases and Exposure Assessment to Brucellosis within rural and peri-urban areas in Kajiado, Kenya. F1000Research, 8, 1916

Pappas, G., Papadimitriou, P., Akritidis, N., Christou, L., Tsianos, E. V. (2006). The new global map of human Brucellosis. Lancet Infect Dis.; 6(2):91–9.

Proch V, Singh B, Schemann K, Gill J, Ward M, Dhand N. 2018. Risk factors for occupational Brucella infection in veterinary personnel in India. Transbound Emerg Dis. 65(3):791–798.

Ragan, V. E. (2021). Infectious agents: brucellosis. Bovine Reproduction, 742–752.

Rubach, M. P., Halliday, J. E., Cleaveland, S., & Crump, J. A. (2013). Brucellosis in low-income and middle-income countries. Current opinion in infectious diseases, 26(5), 404–412.

Salisu, U. S., Kudi, C. A., Bale, J. O. O., Babashani, M., Kaltungo, B. Y., Baba, A. Y., … & Jamilu, Y. R. (2018). Risk factors and knowledge of Brucella infection in camels, attitudes and practices of camel handlers in Katsina State, Nigeria. Nigerian Veterinary Journal, 39(3), 227–239.

Seleem, M. N., Boyle, S. M., & Sriranganathan, N. (2010). Brucellosis: a re-emerging zoonosis. Veterinary microbiology, 140(3-4), 392–398.

Solera, J.; Martínez, A.E.; Espinosa, A.; Castillejos, M.L.; Geijo, P.; Rodríguez-Zapata, M. (1998) Multivariate model for predicting relapse in human brucellosis. J. Infect. 36, 85–92.

Spencer, P. (2012). Nomads in alliance: symbiosis and growth among the Rendille and Samburu of Kenya. Oxford University Press.

Steinfeld, H.; Wassenaar, T.; Jutzi, S. (2006). Livestock production systems in developing countries: Status, drivers, trends. Rev. Sci. Tech. Off. Int. Epiz. 25, 505–516.

TeshomeYimer, B., Feleke, B. E., Bogale, K. A., & Tsegaye, G. W. (2021). Factors Associated with Human Brucellosis among patients Attending in Ayu Primary Hospital, North Showa, Ethiopia: ACase Control Study. Ethiopian Journal of Health Sciences, 31(4), 709–718. 10.4314/ejhs.v31i4.4

Tuon, F. F., Gondolfo, R. B., & Cerchiari, N. (2017). Human-to-human transmission of Brucella–a systematic review. Tropical Medicine & International Health, 22(5), 539–546.

USAID (2020). Effective Engagement with Pastoralist Populations: Guidance for USAID operating units. USAID.

Welburn SC, Beange I, Ducrotoy MJ, Okello AL. The neglected zoonoses—the case for integrated control and advocacy. ClinMicrobiol Infect. 2015;21(5):433–443. 10.1016/j.cmi.2015.04.011

WHO (2015). Neglected Zoonotic Diseases. Available at: https://www.who.int/news-room/facts-in-pictures/detail/neglected-zoonotic-tropical-diseases. Accessed on 24.01/2024.

